# Missed childhood immunizations during the COVID-19 pandemic in Brazil: analyses of routine statistics and of a national household survey

**DOI:** 10.1101/2020.11.30.20240911

**Authors:** Mariangela F Silveira, Cristian T Tonial, Ana Goretti K Maranhão, Antonia MS Teixeira, Pedro C Hallal, Ana Maria B Menezes, Bernardo L Horta, Fernando P Hartwig, Aluisio JD Barros, Cesar G Victora

## Abstract

**Introduction:** There is widespread concern that disruption to health services during the COVID-19 pandemic has led to declines in immunization coverage among young children, but there is limited information on the magnitude of such impact.

**Methods:** We used data from two nationwide sources covering the whole of Brazil. Data from the Information System of the National Immunization Program (SIPNI) on the monthly number of vaccine doses administered to young children were analyzed. The second source was a survey in 133 large cities in the 27 states in the country, carried out from August 24-27. Respondents answered a question on whether children under the age of three years had missed any scheduled vaccinations during the pandemic, and available vaccination cards were photographed for later examination.

**Results:** SIPNI data showed that, relative to January and February 2020, there was a decline of about 20% in vaccines administered to children aged two months or older during March and April, when social distancing was at the highest level in the country. After May, vaccination levels returned to pre-pandemic values. Survey data, based on the interviews and on examination of the vaccine cards, showed that 19.0% (95% CI 17.0;21.1%) and 20.6% (95% CI 19.0;23.1%) of children, respectively, had missed immunizations. Missed doses were most common in the North (Amazon) region and least common in the South and Southeast, and also more common among children from poor than from wealthy families.

**Interpretation:** Our results show that the pandemic was associated with a reduction of about 20% in child vaccinations, but this was reverted in recent months. Children from poor families and from the least developed regions of the country were most affected. There is an urgent need to booster immunization activities in the country to compensate for missed doses, and to reduce geographic and socioeconomic inequalities.

## Introduction

The COVID-19 pandemic has disrupted routine health services throughout the world, due to the need for social distancing and with facilities being affected by staff shortages and overwhelmed with COVID-19 cases.^1^ Specifically in the case of childhood immunizations, campaigns were interrupted and concerns about bringing children to health facilities have contributed to reducing coverage.^2, 3^ Yet, a modeling exercise for Sub-Saharan Africa suggested that child deaths due to vaccine-preventable diseases would far exceed the number of childhood deaths due to COVID-19 illness acquired during visits to health facilities in order to receive vaccines.^4^

Much of the literature on the impact of the pandemic on vaccination coverage consists of modeling exercises with hypothetical data, as few studies are available from low- and middle-income countries that included actual measurements of the shortfall.^5^ A study in Afghanistan showed that due to Covid-19 spread, polio vaccine coverage was reduced and reported cases increased.^6^ At the main university hospital in Saudi Arabia, a retrospective cohort from 2017 to 2020 found that coverage with all childhood immunizations, except for vaccines administered soon after birth were around 20% to 50% lower in 2020 compared with 2017 to 2019.^7^ A study in a province in Pakistan relied upon electronic records to compare immunizations during the lock down period with doses given in the six months preceding the pandemic, and found an average decline of 52.5% in daily doses.^5^ Our literature search failed to identify any studies from Latin America, which has been severely affected by the pandemic.

We analyzed two nationwide data sources to estimate how many children missed immunizations since February 2020, when the first COVID-19 case was reported in Brazil, including the information system for the national immunization system, and a probability survey carried out in August 2020 in 33,250 households from 133 of the largest cities in the country, which included a question on missed vaccinations by young children.

## Methods

### Routine vaccination data

The first data source were routine reports on the number of vaccine doses administered to young children in the country, based on the National Immunization Program Information System (PNI) of the Brazilian Ministry of Health. This open access database^8^ includes data since 1994, being regularly updated by health facilities in the 5,570 municipalities in the country.^9^ Information on vaccine coverage is estimated by dividing reported doses by the number of children in the age range at which the vaccine should have been administered, divided by the numbers of children derived from the National Live Births System.^10^ Information on the following vaccines, all of which are part of the routine vaccine calendar for infants, were obtained:

- Single dose of Bacille Calmette-Guerin (BCG), which is usually administered soon after birth, most often in the maternity wards or during first postnatal visit of the child to a health facility;
- First dose of hepatitis B vaccine, administered within the first 30 days of life, also most often in the maternity hospital or during the first postnatal visit;
- Third dose of pentavalent vaccine (diphtheria, tetanus, pertussis, haemophilus influenza B and hepatitis B), scheduled for administration at the age of six months;
- Third dose of injectable polio vaccine, also scheduled for the age of six months;
- First dose of triple viral vaccine (measles, mumps and rubella) administered at the age of 12 months.

In order to assess the likely impact of the pandemic, we extracted data on the monthly coverage with the above vaccines from 2017 to 2020. We then averaged coverage in 2017 to 2019, and compared with coverage for the same month in 2020 to account for seasonality.^11^

### Nationwide household survey

We used data from the fourth round of the nationwide EPICOVID-19 (http://www.epicovid19brasil.org/) survey, conducted between August 24-27. Brazil is divided into 27 federation Units (26 states and the Brasilia Federal District), which are further divided into 133 intermediary regions. In each of these regions, we selected the most populous city for the study. Within each city, 25 census tracts were chosen with probability proportionate to size, and 10 households were randomly selected per tract based on listings prepared by the Brazilian Institute of Geography and Statistics. Household members were listed using a smartphone app, and for children under the age of three years the survey respondent was asked the following question: “*Did the child miss any scheduled vaccinations since the pandemic started?*” Regardless of the answer, interviewers asked whether the child had an immunization card, and if so, asked the respondent to allow the card to be photographed. The photographs were assessed by two reviewers (MFS and CTT) who classified children according to whether or not they had missed a vaccine dose that was scheduled from March to August 2020.

The questionnaire also provided information on the child’s age and sex, and on the region of the country. Additional information on household assets and characteristics of the building was used in principal component analyses to create a household wealth score, which was later divided into quintiles.^12, 13^

As the primary purpose of the survey was to assess the presence of antibodies against SARS-CoV-2 using a rapid point-of-care test, Interviewers wore individual protection equipment (aprons, gloves, surgical face masks, and shoe and hair covers) that were discarded as hospital waste after each interview. They were tested for COVID-19 prior to the field work, and every two days thereafter. Ethical approval was granted by Brazil’s National Ethics Committee (process number CAAE 30721520.7.1001.5313), with written informed consent from all participants; for children and adolescents, consent was sought from parents or guardians.

Chi-squared tests were used to compare the proportions of children who missed vaccinations according to region of the country, sex, age and household wealth quintiles. For the latter, tests for linear trend in proportions were used. Data analyses took into account the complex nature of the cluster sampling. Additional information on the study is available in its website (www.epidcovid19brasil.org) and in an earlier publication.^14^

## Results

### Routine vaccination data

Figure 1 shows the evolution of vaccine coverage by year from 2017 to the first six months of 2020. Coverage levels were similar in 2017 and 2018, but for all vaccines except MMR coverage was reduced in 2019. In 2020, coverage levels were substantially lower than in the previous years.

**Figure 1.**
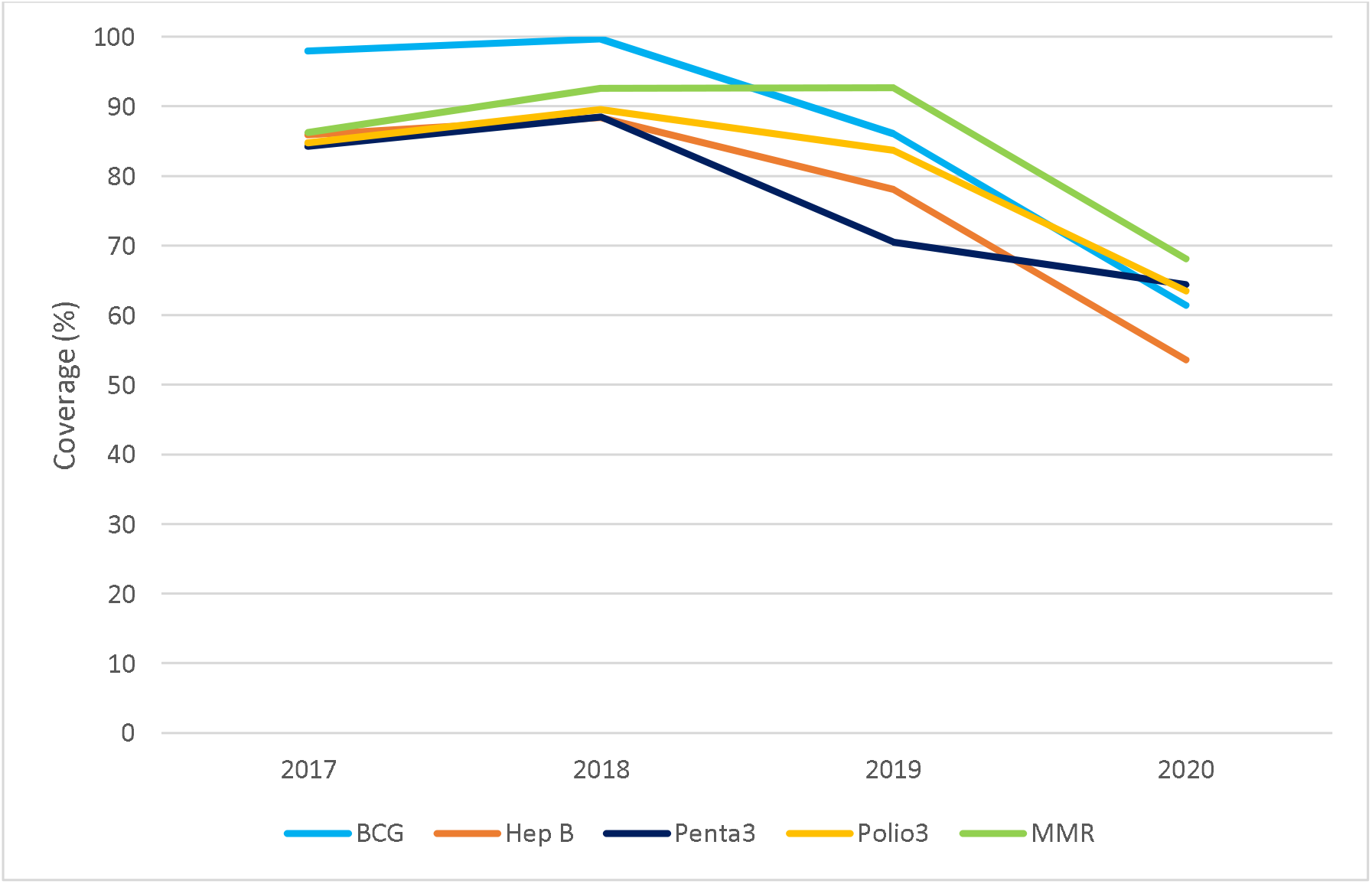
Coverage levels with five vaccines by year, 2017-2020. Source: SI-PNI.

In Figure 2, coverage levels by month in 2020 are shown as ratios relative to average coverage in the corresponding month from 2017-2019. For BCG and hepatitis B vaccines, coverage ratios in 2020 were stable over the six months, at around 63% to 72% of the 2017-2019 values. For the other three vaccines, coverage ratios were considerably lower in March and April than in earlier or later months; in June, levels had returned to almost the same levels as in the previous years, particularly for pentavalent vaccine.

**Figure 2.**
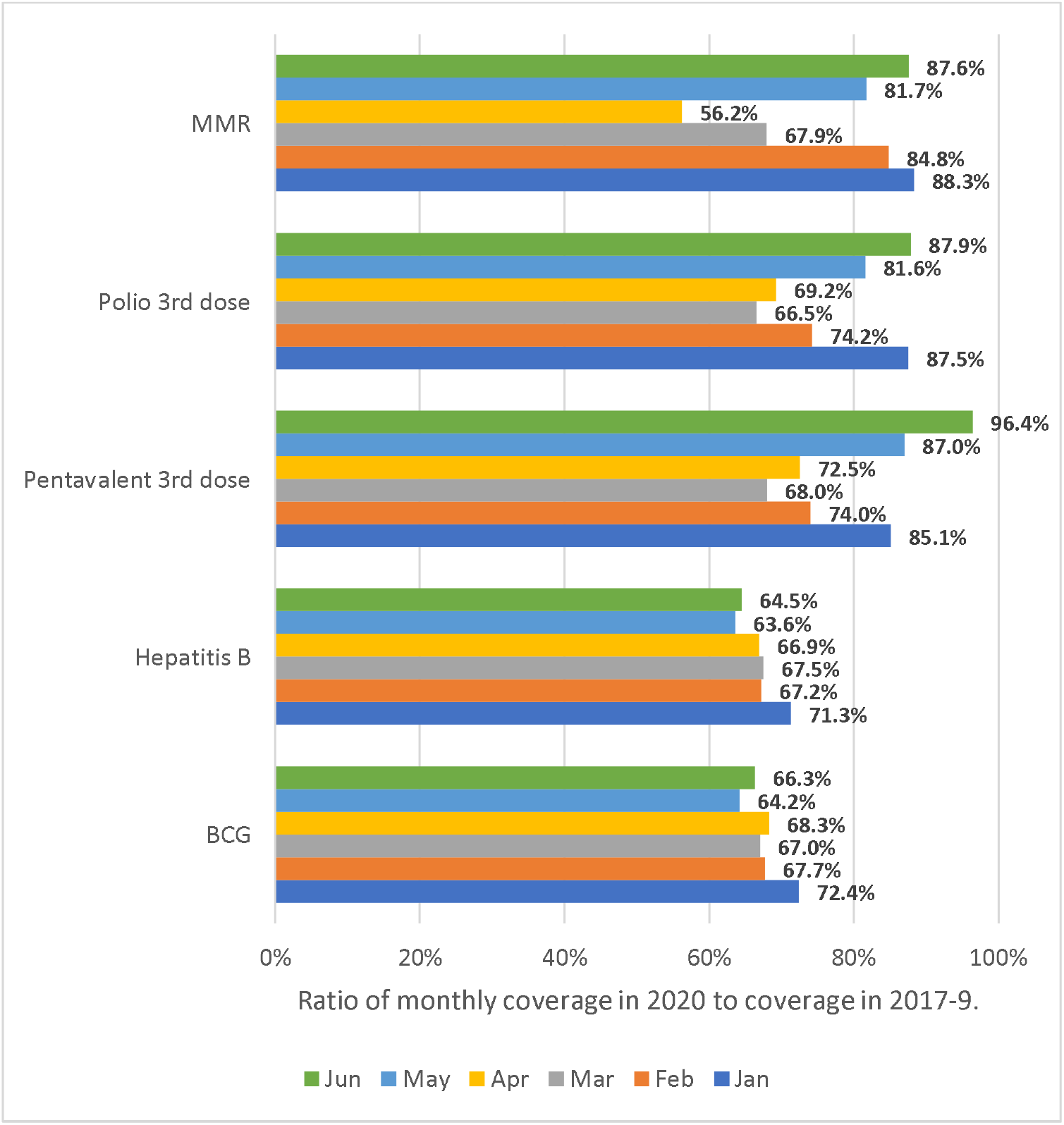
Ratio of monthly coverage in 2020 compared to the same month in 2017-2019. Source: SI-PNI.

**Figure 2.**
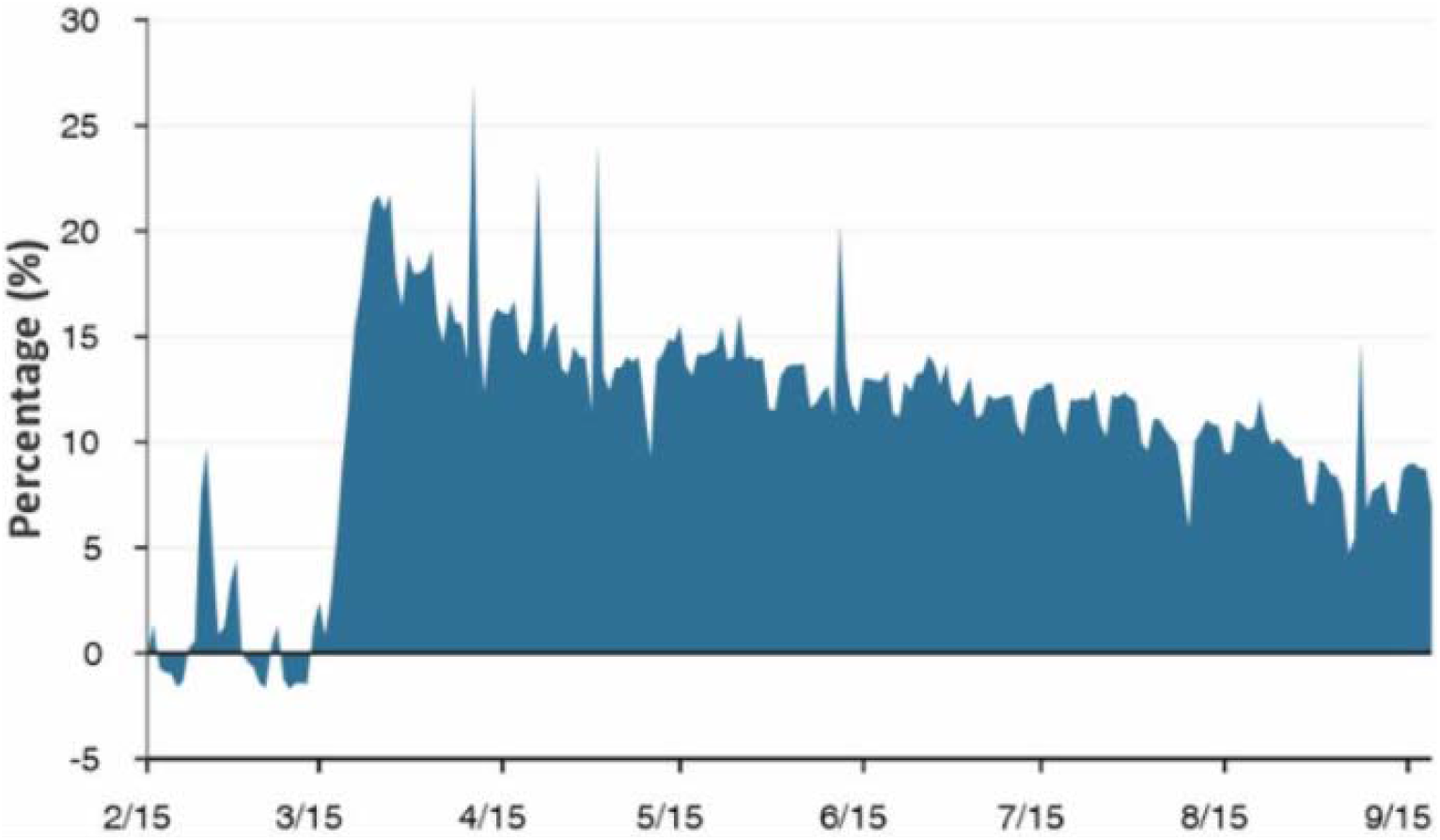
Daily percent changes in residential mobility in Brazil during 2020, compared to the baseline period before the COVID-19 pandemic. Source: Google COVID-19 Community Mobility Reports.

The average change in April and May, compared to the mean values for the other for months, were 0% for BCG, +1% for hepatitis B, -18% for pentavalent and polio, and -27% for MMR.

Figure 2 shows that the implementation of social distancing was strongest in the months of March and April, which were the months with the most marked drops in immunizations.

### Nationwide household survey

The overall response rate in the nationwide survey was 55%, due to logistic difficulties during lockdown period. No family members were at home in 22% of the households, and in another 23% the residents refused to undergo a test for COVID antibodies. Questionnaire information on missed vaccination was obtained for 2,530 children under the age of three years. Of these, it was possible to photograph vaccination cards for 1,785 (69.6%), 22.8% either did not have a card or could not find it, and 7.6% refused to allow the interviewers to photograph the card. Of the 1,785 cards, 1,547 (86.7%) yielded where vaccination dates were readable.

The proportions of children who missed scheduled vaccinations were 19.0% (95% CI 17.0;21.1%) and 20.6% (95% CI 19.0;23.1%), respectively. Agreement between the two indicators was equal to 72.3%. Table 1 shows the proportions of children who missed vaccinations by region, sex, age and household wealth. Results from both methods were quite similar. The highest proportions of default were in the North and Center-West regions, and the lowest in the South. There were no significant differences by sex. Infants were less likely to miss vaccinations than one-year-old children but results for two-year-old children were inconsistent, with extremely low frequency according to the vaccination card. Patterns according to household wealth were very consistent, showing that children from poor families were more likely to have missed vaccine doses.

**Table 1.** Proportions of children who missed scheduled vaccinations according to the questionnaire and vaccine card, stratified by region, sex, age and wealth quintiles.

|  |  | Questionnaire |  | Vaccine card |  |
| --- | --- | --- | --- | --- | --- |
|  |  | Number | Missed vaccines (%) | Number | Missed vaccines (%) |
| **Region** | North | 542 | 25.7<br>(22.0; 29.7) | 359 | 28.4<br>(24.2; 33.1) |
|  | Northeast | 789 | 19.4<br>(16.8; 22.3) | 506 | 18.4<br>(15.2; 22.1) |
|  | Southeast | 524 | 15.1<br>(12.1; 18.6) | 320 | 16.6<br>(12.8; 21.2) |
|  | South | 332 | 12.7<br>(9.6; 16.5) | 199 | 16.1<br>(11.7; 21.7) |
|  | Center-west | 252 | 20.6<br>(16.0; 26.1) | 158 | 27.2<br>(21.0; 34.5) |
|  |  |  | *P<0.001* |  | *P<0.001* |
| **Sex** | Boys | 1259 | 20.2<br>(18.0; 22.5) | 810 | 21.0<br>(18.3; 24.0) |
|  | Girls | 1180 | 17.9<br>(15.8; 20.2) | 732 | 20.9<br>(18.1; 24.0) |
|  |  |  | *P=0.14* |  | *P=0.97* |
| **Age (years)** | 0 | 493 | 15.4<br>(12.5; 18.9) | 363 | 25.6<br>(21.4; 30.4) |
|  | 1 | 939 | 20.1<br>(17.6; 22.9) | 583 | 35.0<br>(31.2; 39.0) |
|  | 2 | 1007 | 19.9<br>(17.5; 22.4) | 601 | 4.5<br>(3.1; 6.6) |
|  |  |  | *P=0.05* |  | *p<0.001* |
| **Wealth quintile** | Poorest | 639 | 22.5<br>(19.3; 26.2) | 421 | 24.5<br>(20.6; 28.8) |
|  | 2nd | 514 | 21.0<br>(17.7; 24.7) | 318 | 24.5<br>(20.2; 29.4) |
|  | 3rd | 489 | 17.0<br>(13.9; 20.6) | 316 | 19.0<br>(15.0; 23.7) |
|  | 4th | 429 | 17.5<br>(14.0; 21.6) | 269 | 17.8<br>(13.6; 23.0) |
|  | Wealthiest | 368 | 15.0<br>(11.6; 19.1) | 218 | 15.6<br>(11.3; 21.1) |
|  |  |  | *P=0.03* |  | *P=0.01* |
| **Total** |  | 2439 | 19.0<br>(17.0; 21.1) | 1547 | 21.0<br>(19.0; 23.1) |

## Discussion

The Brazilian National Immunization Program (PNI) was created in 1973, and over time the country became one of the most successful programs in the world with coverage levels close to 100% for most vaccines by the beginning of the century.^15^ The last polio case was documented in 1989 and the disease was officially recognized as eradicated in 1994, and in 2016 measles was also certified as eradicated in Latin America as a whole.^16^ The immunization program incorporated new vaccines over time. At present, a child must be brought to a health facility on seven different occasions during the first year of life to receive 15 different doses of vaccines against 11 infectious diseases. The growing complexity of the vaccination schedule requiring multiple visits to health facilities, associated with a general perception that vaccine-preventable diseases are no longer a risk for children,^9^ as well as the emergence of vaccine hesitancy^17^ have contributed to substantial declines in vaccine coverage throughout the country in recent years.^9, 18^ Our own results on annual coverage confirm that a decline was already evident in 2019, before the pandemic (Figure 1). Drops in coverage led to a reversal in the country’s measles-free status. This disease was reintroduced from Venezuela in 2018 and as a consequence of declining coverage levels, most states in the country have reported measles cases since then.^19^ It is against this background scenario of declining coverage that the impact of the COVID-19 pandemic must be interpreted.

The present results, based upon two different data sources, confirm the negative impact of the pandemic on child vaccination coverage. The routine system showed a time-delimited drop during the months of March and April. Data from Google residential mobility (Figure 3) confirm that these were the two months in 2020 where the population was most likely to remain isolated at their homes. Starting in May, isolation was gradually relaxed. An important findings is that BCG and the first does for hepatitis B, which are often administered in the maternity hospital or in the first postnatal visit to a facility, were not affected by the pandemic, whereas vaccines that administered to older children showed a clear dip in March and April followed by a recovery from May onwards. The magnitude of the decline was of 18% for pentavalent and polio vaccines, and 27% for MMR vaccine. A possible explanation for this difference is virtually all children in the country are born in a hospital, and even if BCG or hepatitis B vaccines are not administered in the maternity ward, families are more motivated to attend the first postnatal check-up than to bring older children for immunization appointments, which may be perceived as less urgent.

Results from the national household survey, based both on questionnaire responses and inspection of vaccine cards, suggest that about 20% of children failed scheduled vaccination appointments since March. According to both methods, missed vaccinations were most common in the North region and among children from the poorest families. The main discrepancy was in missed appointments for two-year-old children, which were uncommon according to vaccination cards, but frequent according to the questionnaire. For children who are up to date with their vaccinations, no doses are scheduled for the third year of life, but for those who are not up to date, health workers will schedule visits during the year to delivered the vaccines that had been missed in previous years. It is possible that questionnaires respondents – usually the children’s parents – were confused about the need for immunizations in the third year. The social patterning of missed doses is not unexpected, as coverage typically increases with family wealth in most low- and middle-income countries.^20, 21^ Similar inequality patterns have been reported by Brazilian researchers, ^22, 23^, although vaccine hesitancy seems to be rising among better-off families in the wealthiest regions of the country.^17^ Higher frequencies of missed doses among the poor may be perhaps be explained by characteristics of health facilities (geographic distribution, staffing, opening hours), to concerns about infection by bringing children to crowded services, and to the need to rely on public transportation with consequent exposure to the virus. In contrast, better off families are more likely to rely on private transportation and to also have access to private clinics.

Our analyses have limitations. The definitions of missed vaccinations based on the national information system and on the household survey are different, yet both sources suggest that about one in five children missed vaccinations. Coverage of the national information system is high, but there are delays in reporting from primary care facilities; nevertheless, it is unlikely that delays can explain the present findings of a clear dip during two calendar months and return to previous levels afterwards, as well as the finding that vaccines delivered primarily in hospitals were not affected. The response rate in the national survey was low, at around 55%, which is explained by the fact that many homes were empty as families attempted to move away from large cities due to the pandemic, as well as by refusals to receive interviewers due to fear of infection. Although the sample included all regional hub cities in the country, it is not representative of smaller towns and rural areas, where vaccine coverage may be different.

On the other hand, the strengths of our analyses include the use of two different data sources that provided consistent results on levels and patterns of missed vaccinations. Due to the national scope of our study in a country with 220,000,000 inhabitants, it represents the largest assessment so far on how the COVID-19 pandemic has affected immunizations in a low- or middle-income country.

Although the national data suggest that vaccine administration has returned to pre-pandemic levels, the dip of about 20% observed during March and April has not yet been compensated by higher vaccination rates in recent year. Also of concern is that the impact of the pandemic has been larger among children from poor families in the poorest regions of the country – who are most likely to die from infectious diseases^24^ - thus further accentuating social inequalities in coverage that were already present in the recent past. There is a strong need to reinvigorate the national immunization program to tackle the impact of the pandemic.

## Data Availability

Data is available at http://www.epicovid19brasil.org/?page_id=472

## Acknowledgments

The study was funded by the Brazilian Ministry of Health, Instituto Serrapilheira, Brazilian Collective Health Association (ABRASCO) and the JBS S.A. initiative ‘Fazer o Bem Faz Bem’.

## Author’s contributions

Mariângela F Silveira, Cristian T Tonial, Ana Goretti K Maranhão, Antonia MS Teixeira, Pedro C Hallal, Ana M B Menezes, Bernardo L Horta, Fernando P Hartwig, Aluísio J D Barros, and Cesar G Victora contributed to the conception and design of the work, to the acquisition, analysis, and interpretation of data and the draft of the manuscript.

